# Prevalence of Chronic Diseases, Depression, and Stress among U.S. Child Care Professionals during the COVID-19 Pandemic

**DOI:** 10.1101/2022.03.01.22271717

**Authors:** Jad A. Elharake, Mehr Shafiq, Ayse Cobanoglu, Amyn A. Malik, Madeline Klotz, John Eric Humphries, Thomas Murray, Kavin M. Patel, David Wilkinson, Inci Yildirim, Rachel Diaz, Rosalia Rojas, Anael Kuperwajs Cohen, Aiden Lee, Chin R. Reyes, Saad B. Omer, Walter S. Gilliam

## Abstract

**Importance:** There is no published national research reporting child care professionals’ physical health, depression, or stress during the COVID-19 pandemic. Given their central role in supporting children’s development, child care professionals’ overall physical and mental health is important.

**Objectives:** To evaluate the prevalence of chronic diseases, depression, and stress levels during the COVID-19 pandemic among U.S. child care professionals.

**Design:** In this large-scale national survey, data were collected through an online survey from May 22, 2020 to June 8, 2020. We analyzed the association of sociodemographic characteristics with four physical health conditions (asthma, heart disease, diabetes, and obesity), depression, and stress weighted to national representativeness. Sociodemographic characteristics included race, ethnicity, age, gender, medical insurance status, and child care type.

**Setting:** Center- and home-based child care.

**Participants:** Child care professionals (n = 81,682) from all U.S. states and the District of Columbia.

**Results:** Mean age was 42.1 years (standard deviation = 14.1); 96.0% (n = 78,725) were female, 2.5% (n = 2,033) were male, and 0.3% (n = 225) were non-binary. For physical health conditions, 14.3% (n = 11,717) reported moderate to severe asthma, 6.5% (n = 5,317) diabetes, 4.9% (n = 3,971) heart disease, and 19.8% (n = 16,207) being obese. Regarding mental health, 45.7% (n = 37,376) screened positive for depression and 66.5% (n = 54,381) reported moderate to high stress levels. Race, ethnicity, and gender disparities were evidenced for physical health conditions of child care professionals, but not for mental health during the pandemic.

**Conclusions and Relevance:** Our findings highlight that child care professionals’ depression rates during the pandemic were much higher than before the pandemic, and depression, stress and asthma rates were higher than U.S. adult depression rates during the pandemic. Given the essential work child care professionals provide during the pandemic, policy makers and public health officials should consider what can be done to support the physical and mental health of child care professionals.

**Key Points:** *Question:* What is the prevalence of chronic diseases, depression, and stress among U.S. child care professionals during the COVID-19 pandemic?

*Findings:* In this survey of 81,682 U.S. child care professionals, 14.3% reported moderate to severe asthma, 6.5% diabetes, 4.9% heart disease, 19.8% being obese, 45.7% screening positive for depression, and 66.5% moderate to high stress levels.

*Meaning:* During the pandemic, child care professionals reported depression rates much higher than before the pandemic, and asthma, stress, and depression much greater than U.S. adult estimates, highlighting a need for effective supports for the wellbeing of this essential workforce.

## Introduction

There are approximately 1.1 million paid and registered child care professionals in the United States (U.S.) providing care for 10 million children across center- and home-based settings.^1,2^ While child care professionals play a central role in children’s learning and personal development, they represent a highly vulnerable workforce,^3^ consisting largely of women and in many child care sectors racial and ethnic minorities, immigrants, and lower-income individuals.^4^ As an historically exploited labor group,^5^ child care is one of the lowest-paid occupations in the U.S., with an annual salary 67% below the national average.^3^ Child care professionals face challenging work conditions including long hours and high job demands,^6-9^ leading to staff turnover, absenteeism, poor physical health conditions, high rates of burnout, emotional exhaustion, and mental health.^10^

The unpredictable nature and impact of the coronavirus disease-2019 (COVID-19) pandemic has further exacerbated the financial and work-related stress faced by child care professionals.^11,12^ Despite extensive mitigation strategies to contain the pandemic, many child care programs closed in March 2020.^13^ Unlike many schools and universities that fully transitioned to online learning, there were no alternative virtual platforms for a majority of child care programs.^13^ As a result, more than 35% of jobs in the child care industry were lost.^14^ Not only did this threaten the financial stability of child care professionals and families that rely on child care for their employability,^1,15,16^ it also limited children’s educational, social, and nutritional opportunities^17^ and may have further worsened the physical and mental health of child care professionals.^11,12^

There is no published national research reporting child care professionals’ physical health, depression, or stress during the COVID-19 pandemic. Pre-pandemic studies on child care professionals have reported a 7.3% rate of diabetes,^18^ obesity rates ranging from 34% to 66.3%,^18-21^ and clinical depression rates ranging from 16.0% to 36.1%.^21-25^ These rates compare to the overall rates for U.S. adult women of 8.7% for diabetes,^26^ 41.9% for obesity,^27^ 9.6% for depression pre-pandemic,^28^ and 27.8% to 32.8% for depression during the pandemic.^29-31^ There is only one study focusing on the stress levels of child care professionals during the pandemic, conducted in Indiana, which found that 63% of child care professionals reported moderate to high stress levels,^12^ almost twice the national estimate of 37% for U.S. adults during the pandemic.^32^ Prior to the pandemic, 36.8% to 62.1% of child care professionals were reported to be experiencing moderate to high levels of stress.^12,20,33^

Given that the well-being of child care professionals is associated with children’s academic and emotional learning outcomes,^9,25,33,34^ it is imperative to understand and address the condition of child care professionals during the pandemic and beyond. While relatively small-scale studies have documented the mental health status of early childhood educators prior to the COVID-19 pandemic,^9,18-21,24,25,33,35-37^ very little is known about their physical and mental health status during the pandemic. This study evaluates the prevalence of chronic diseases, depression, and stress levels during the COVID-19 pandemic among U.S. child care professionals.

## Methods

### Study Design and Participants

Data were collected from self-identifying child care professionals through an online Qualtrics® survey distributed from May 22, 2020 to June 8, 2020, about 10 to 13 weeks into the COVID-19 pandemic, through various contact lists of individuals associated with the child care industry, as described previously.^38^ Inclusion criteria were participants who self-identified as child care professionals working in child care before or during the pandemic; consented to the study; were 18 years or older; and resided in a U.S. state or the District of Columbia. Of the 94,390 individuals who accessed the survey, 82,613 satisfied inclusion criteria and 81,682 (98.9%) provided data necessary for analyses. Participants were offered entry into a raffle for one of 20 $500 gift cards. The research protocol was approved by the Yale University Institutional Review Board as a Category 2(ii) exempt protocol (#2000028232).

### Variables

#### Chronic Diseases and Health Conditions

Ten chronic diseases and physical health conditions, identified as risk factors for COVID-19 complications by the Centers for Disease Control and Prevention (CDC) at the time of the survey,^39^ were asked in the survey with respondents indicating which apply to them: chronic lung disease/chronic obstructive pulmonary disease (COPD), chronic/severe kidney disease, liver disease, heart disease, immune-compromising conditions (such as immune deficiencies bone marrow/organ transplant), immunosuppressive treatments for cancer or an inflammatory disease (such as lupus, rheumatoid arthritis, etc.), smoking, diabetes, asthma (moderate to severe), and obesity. Asthma, diabetes, heart disease, and obesity had the highest prevalence in our sample and were therefore selected for further analysis. Prevalence of the remaining six physical health conditions were analyzed by sociodemographic characteristics.

#### Depressive Symptoms

The 10-item Center for Epidemiological Studies–Depression (CES-D-10) scale is a reliable and valid self-report scale designed to measure depressive symptomatology and screen for Major Depression.^40^ Items assess depression-related symptoms experienced in the past week (0 = Rarely or none of the time; 1 = Some or little of the time; 2 = Occasionally or a moderate amount of the time; 3 = All of the time), such as restless sleep, poor appetite, and feelings of loneliness, with positively stated items reverse coded prior to calculating sum scores. As in other studies,^30,41,42^ sum scores greater than or equal to 10 were considered positive for depression.

#### Stress

The 10-item Perceived Stress Scale (PSS-10) is a validated short-form version of the Perceived Stress Scale, the most widely used psychological instrument for measuring the perception of stress.^43^ Questions ask about feelings and thoughts during the last month and are rated on a five-point Likert Scale (0 = Never; 1 = Almost Never; 2 = Sometimes; 3 = Fairly Often; 4 = Very Often).^44^ Positively stated items were reverse coded. Sum scores ranging from 0-13 are considered low stress, 14-26 are moderate stress, and 27-40 are high stress.^45^

#### Sociodemographic Factors

Respondents were asked to provide information on their racial and ethnic backgrounds with items worded identically to the most recent U.S. Census questionnaire. Options for race included White, Black/African American, American Indian/Alaska Native, Native Hawaiian/Pacific Islander, and Asian, and respondents who selected more than one race were coded as ‘multiracial.’ Additionally, ethnicity (Hispanic, Latino, or Spanish Origin versus not), access to medical insurance, gender, age, and child care program type were considered in the analysis.

### Data Weighting and Missing Data Analysis and Treatment

The sample was weighted to national representativeness for U.S. child care professionals by state, age, race, and ethnicity based on the 2019 American Community Survey,^46^ with the top and bottom 5% of the weights trimmed to reduce sampling variance. Missing data analysis and multiple imputation of missing data are described in **eMethods**.

### Statistical Analysis

Descriptive statistics were used to present all variables. Separate multivariable logistic regression models were used to assess the association of covariates (age, race, ethnicity, gender, program type, and medical insurance) with prevalence of heart disease, asthma, diabetes, and obesity, individually. Separate multivariable linear regression models were used to assess the association of these covariates with self-reported depression and stress scores. Significance was set to alpha = 0.05, two-tailed. Analyses were conducted in SPSS Statistics (version 28.0.1.) and R (version R.4.1.1.).

## Results

### Demographics Characteristics

Of the total sample (n = 81,682), the mean age was 42.1 years (standard deviation [std] = 14.1). Across racial categories, 63.8% (n = 52,164) were White, 14.5% (n = 11,837) Black/African American, 3.6% (n = 2,949) Asian, 3.6% (n = 2,944) multiracial, 1.9% (n = 1,582) American Indian/Alaska Native, 0.6% (n = 491) Native Hawaiian/Pacific Islander, and 11.9% (n = 9,731) preferred not to identify their race. Across the ethnic category, 21.7% (n = 17,753) identified as Hispanic. Also, 96.4% (n = 78,725) of the sample identified as female, 2.5% (n = 2,033) as male, and 0.3% (n = 225) as non-binary. Most respondents worked in child care centers, with 24.5% (n = 19,976) in for-profit centers. Lastly, 89.2% (n = 72,890) of the sample reported access to medical insurance (**Table 1**).

**Table 1:** Sample Demographics and Prevalence of Physical Health Conditions by Sociodemographic Characteristics.

| Characteristic | Physical Health Conditions |  |  |  |  |
| --- | --- | --- | --- | --- | --- |
|  | Total<br>N (%) | Asthma<br>N (%) | Heart Disease<br>N (%) | Diabetes<br>N (%) | Obesity<br>N (%) |
| **Total** | 81682 | 11717 (14.3) | 3971 (4.9) | 5317 (6.5) | 16207 (19.8) |
| **Age (Mean [std])** | 42.1 (14.1) | 40.9 (14.1) | 50.3 (13.8) | 51.3 (12.8) | 44.0 (13.3) |
| **Race** |  |  |  |  |  |
| **White** | 52164 (63.8) | 7413 (14.2) | 2520 (4.8) | 2946 (5.6) | 10905 (20.9) |
| **American Indian/Alaska Native** | 1582 (1.9) | 298 (18.8) | 91 (5.8) | 178 (11.2) | 330 (20.9) |
| **Asian** | 2949 (3.6) | 325 (11.0) | 121 (4.1) | 195 (6.6) | 195 (6.6) |
| **Black/African American** | 11837 (14.5) | 1787 (15.1) | 674 (5.7) | 1209 (10.2) | 2587 (21.9) |
| **Multiracial** | 2944 (3.6) | 638 (21.7) | 179 (6.1) | 130 (4.4) | 703 (23.9) |
| **Native Hawaiian/Pacific Islander** | 491 (0.6) | 67 (13.6) | 21 (4.3) | 53 (10.9) | 77 (15.7) |
| **Prefer to not answer** | 9731 (11.9) | 1189 (12.2) | 364 (3.7) | 606 (6.2) | 1410 (14.5) |
| **Hispanic** |  |  |  |  |  |
| **No** | 61806 (75.7) | 9099 (14.7) | 3207 (5.2) | 4043 (6.5) | 13121 (21.2) |
| **Yes** | 17753 (21.7) | 2362 (13.3) | 676 (3.8) | 1112 (6.3) | 2723 (15.3) |
| **Prefer to not answer** | 2139 (2.6) | 255 (11.9) | 88 (4.1) | 163 (7.6) | 363 (17.0) |
| **Gender** |  |  |  |  |  |
| **Female** | 78725 (96.4) | 11358 (14.4) | 3789 (4.8) | 5106 (6.5) | 15780 (20.0) |
| **Male** | 2033 (2.5) | 231 (11.4) | 139 (6.8) | 149 (7.3) | 267 (13.1) |
| **Non-Binary** | 225 (0.3) | 51 (22.8) | 14 (6.3) | 11 (4.8) | 51 (22.7) |
| **Prefer to not answer** | 715 (0.9) | 76 (10.6) | 29 (4.1) | 52 (7.3) | 109 (15.2) |
| **Insurance** |  |  |  |  |  |
| **No** | 8808 (10.8) | 1012 (11.5) | 327 (3.7) | 424 (4.8) | 1551 (17.6) |
| **Yes** | 72890 (89.2) | 10705 (14.7) | 3644 (5.0) | 4893 (6.7) | 14656 (20.1) |
| **Program Type** |  |  |  |  |  |
| **Center-based** |  |  |  |  |  |
| **For-profit center** | 19976 (24.5) | 2861 (14.3) | 1050 (5.3) | 1222 (6.1) | 4133 (20.7) |
| **School-based** | 10604 (13.0) | 1529 (14.4) | 450 (4.2) | 465 (4.4) | 1729 (16.3) |
| **Head Start / Early Head Start** | 8506 (10.4) | 1309 (15.4) | 370 (4.4) | 637 (7.5) | 1992 (23.4) |
| **Drop-in center** | 1596 (2.0) | 220 (13.8) | 49 (3.1) | 69 (4.3) | 210 (13.1) |
| **Not-for-profit agency center** | 15875 (19.4) | 2336 (14.7) | 802 (5.1) | 1076 (6.8) | 3588 (22.6) |
| **Other center-based** | 5949 (7.3) | 945 (15.9) | 267 (4.5) | 352 (5.9) | 1073 (18.0) |
| **Home-based / Family Child Care** | 18078 (22.1) | 2348 (13.0) | 946 (5.2) | 1435 (7.9) | 3309 (18.3) |
| **Nanny / In-Home Child Care** | 1115 (1.4) | 169 (15.2) | 37 (3.3) | 61 (5.4) | 174 (15.6) |

**Legend:** The total column represents numbers and percentages of sociodemographic characteristics that make up the total sample. Diabetes, Heart Disease, Asthma, and Obesity columns show the numbers and percentages of the prevalence of the disease in that sociodemographic group (i.e., how many participants in a certain group reported having the disease). Due to sample weighting, frequencies may not add exactly to totals.

### Chronic Diseases and Physical Health Conditions

Of the 10 chronic diseases and physical health conditions considered, the greatest rates were found for asthma (14.3%; n = 11,717), diabetes (6.5%; n = 5,317), heart disease (4.9%; 3,971), and obesity (19.8%; n = 16,207). Prevalence for each across sociodemographic characteristics can be found in **Table 1**. Significant sociodemographic predictors for each are presented below and full results are presented in **Table 2**.

**Table 2:** Logistic Regression of Physical Health Conditions and Sociodemographic Characteristics.

| Characteristic | Asthma<br>OR (95% CI) | Heart Disease<br>OR (95% CI) | Diabetes<br>OR (95% CI) | Obesity<br>OR (95% CI) |
| --- | --- | --- | --- | --- |
| Age | **0.99 (0.99 to 0.99)** | **1.04 (1.04 to 1.05)** | **1.05 (1.05 to 1.06)** | **1.01 (1.01 to 1.01)** |
| Race |  |  |  |  |
| White |  | *Reference* |  |  |
| American Indian/Alaska Native | **1.44 (1.23 to 1.69)** | **1.42 (1.11 to 1.82)** | **2.34 (1.96 to 2.80)** | 1.11 (0.95 to 1.29) |
| Asian | **0.74 (0.64 to 0.84)** | 0.89 (0.72 to 1.11) | **1.34 (1.11 to 1.62)** | **0.27 (0.22 to 0.31)** |
| Black/African American | **1.09 (1.02 to 1.16)** | **1.14 (1.03 to 1.26)** | **1.86 (1.71 to 2.03)** | 1.01 (0.96 to 1.07) |
| Multiracial | **1.62 (1.47 to 1.79)** | **1.74 (1.46 to 2.08)** | 1.07 (0.87 to 1.31) | **1.30 (1.18 to 1.43)** |
| Native Hawaiian/Pacific Islander | 1.00 (0.73 to 1.37) | 0.88 (0.53 to 1.45) | **1.88 (1.34 to 2.64)** | **0.76 (0.58 to 0.99)** |
| Prefer to not answer | 0.91 (0.83 to 0.99) | 0.95 (0.81 to 1.11) | 1.09 (0.96 to 1.23) | **0.78 (0.72 to 0.85)** |
| Hispanic |  |  |  |  |
| No |  | *Reference* |  |  |
| Yes | **0.90 (0.85 to 0.96)** | 0.95 (0.85 to 1.06) | **1.34 (1.22 to 1.47)** | **0.75 (0.71 to 0.80)** |
| Prefer to not answer | 0.90 (0.76 to 1.07) | 0.82 (0.61 to 1.12) | 1.23 (0.98 to 1.54) | 0.93 (0.80 to 1.08) |
| Gender |  |  |  |  |
| Female |  | *Reference* |  |  |
| Male | **0.74 (0.63 to 0.88)** | **1.49 (1.22 to 1.80)** | 1.18 (0.96 to 1.44) | **0.62 (0.53 to 0.73)** |
| Non-Binary | **1.58 (1.11 to 2.25)** | **1.97 (1.01 to 3.84)** | 1.06 (0.45 to 2.48) | 1.32 (0.90 to 1.93) |
| Prefer to not answer | 0.80 (0.55 to 1.17) | 0.93 (0.53 to 1.65) | 0.99 (0.66 to 1.50) | 0.82 (0.63 to 1.07) |
| Insurance |  |  |  |  |
| Yes |  | *Reference* |  |  |
| No | **0.74 (0.68 to 0.80)** | 0.89 (0.77 to 1.02) | **0.81 (0.72 to 0.92)** | **0.94 (0.88 to 1.00)** |
| Program Type |  |  |  |  |
| For-profit center |  | *Reference* |  |  |
| Home-based | 0.94 (0.88 to 1.01) | **0.83 (0.74 to 0.92)** | 0.99 (0.89 to 1.10) | **0.85 (0.80 to 0.91)** |
| Nanny/home-visiting | 1.02 (0.84 to 1.25) | 0.89 (0.58 to 1.36) | 1.27 (0.91 to 1.77) | **0.82 (0.67 to 1.00)** |
| Not for-profit center | 1.05 (0.98 to 1.12) | **0.87 (0.78 to 0.97)** | 1.01 (0.91 to 1.11) | **1.10 (1.03 to 1.17)** |
| School-based | 0.99 (0.91 to 1.08) | 0.90 (0.78 to 1.04) | **0.78 (0.69 to 0.88)** | **0.81 (0.75 to 0.87)** |
| Head Start / Early Head Start | 1.08 (0.99 to 1.17) | 0.89 (0.77 to 1.02) | **1.21 (1.08 to 1.35)** | **1.28 (1.19 to 1.37)** |
| Drop-in center | 0.92 (0.78 to 1.09) | 0.86 (0.59 to 1.24) | 0.95 (0.69 to 1.30) | **0.69 (0.57 to 0.84)** |
| Other center-based | **1.14 (1.03 to 1.26)** | 0.91 (0.77 to 1.06) | 1.00 (0.87 to 1.16) | **0.89 (0.81 to 0.98)** |

#### Asthma

Compared to White child care professionals, American Indian/Alaska Native (Odds ratio [OR] = 1.44; 95% Confidence Interval [CI] = 1.23-1.69), Black/African American (OR = 1.09; 95% CI = 1.02-1.16), and multiracial (OR = 1.62; 95% CI = 1.47-1.79) child care professionals had higher odds of having asthma, while controlling for other covariates, whereas Asian (OR = 0.74; 95% CI = 0.64-0.84) participants had lower odds of having asthma. Participants who identified as Hispanic had lower odds (OR = 0.90; 95% CI = 0.85-0.96) of having asthma than those who did not, while controlling for other covariates. Compared to females, participants who identified as non-binary had higher odds of asthma (OR = 1.58; 95% CI = 1.11-2.25). Those who did not have medical insurance had lower odds of having asthma (OR = 0.74; 95% CI = 0.68-0.80) than those who did have medical insurance, while controlling for other covariates.

#### Diabetes

Compared to White child care professionals, American Indian/Alaska Native (OR = 2.34; 95% CI = 1.96-2.80), Asian (OR = 1.34; 95% CI = 1.11-1.62), Black/African American (OR = 1.86; 95% CI = 1.71-2.03), and Native Hawaiian/Pacific Islander (OR = 1.88; 95% CI = 1.34-2.64) child care professionals had higher odds of having diabetes, while controlling for other covariates. Participants who identified as Hispanic had higher odds (OR = 1.34; 95% CI = 1.22-1.47) of having diabetes than those who did not, while controlling for other covariates. Those who did not have medical insurance had lower odds of having diabetes (OR = 0.81; 95% CI = 0.72-0.92) than those who did have medical insurance, while controlling for other covariates.

#### Heart Disease

Compared to White child care professionals, American Indian/Alaska Native (OR = 1.42; 95% CI = 1.11-1.82), Black/African American (OR = 1.14; 95% CI = 1.03-1.26), and multiracial (OR = 1.74; 95% CI = 1.46-2.08) child care professionals had higher odds of having heart disease, while controlling for ethnicity, gender, age, insurance status, and program type. Males (OR = 1.49; 95% CI = 1.22-1.80) and non-binary participants (OR = 1.97; 95% CI = 1.01-3.84) had higher odds of having heart disease than did females.

#### Obesity

Compared to White child care professionals, multiracial (OR = 1.30; 95% CI = 1.18-1.43) child care professionals had higher odds of having obesity, while controlling for other covariates, whereas Asian (OR = 0.27; 95% CI = 0.22-0.31) and Native Hawaiian/Pacific Islander (OR = 0.76; 95% CI = 0.58-0.99) participants had lower odds of having obesity. Participants who identified as Hispanic had lower odds (OR = 0.75; 95% CI = 0.71-0.80) of having obesity than those who did not, while controlling for other covariates. Those who did not have medical insurance had lower odds of having obesity (OR = 0.94; 95% CI = 0.88-1.00) than those who had medical insurance, while controlling for other covariates.

#### Other Physical Health Conditions

Prevalence of the remaining six chronic diseases and physical health conditions were 4.4% (n = 3,619) for smoking, 4.7% (n = 3,851) for immune weakening medications, 2.3% (n = 1,884) for immune compromising conditions, 1.0% (n = 814) for chronic lung disease/COPD, 0.7% (n = 562) for chronic/severe kidney disease, and 0.7% (n = 545) for liver disease. Rates by sociodemographic characteristics are in **eTable 1**. Overall, 26.2% (n = 21,398) of respondents reported at least one medically compromising condition, 9.7% (n = 7,962) reported two, and 4.0% (n = 3,239) reported three or more, as shown in **eTable 2**.

### Mental Health

#### Depressive Symptoms

Of the total sample, 45.7% (n = 37,376) of child care professionals screened positive for depression (CES-D-10 ≥ 10), with a mean score of 10.2 (std = 6.0). The prevalence of clinically relevant depressive symptoms and mean CES-D-10 scores across sociodemographic characteristics can be found in **Table 3**. For every one-year increase in age, on average, the CES-D-10 sum score decreased (β = -0.11; 95% CI = -0.16 to -0.05). Compared to participants who worked in for-profit centers, participants in home-based programs reported lower CES-D-10 sum scores (β = -2.30; 95% CI = -3.89 to -0.72), while controlling for other covariates (**Table 4**).

**Table 3:**
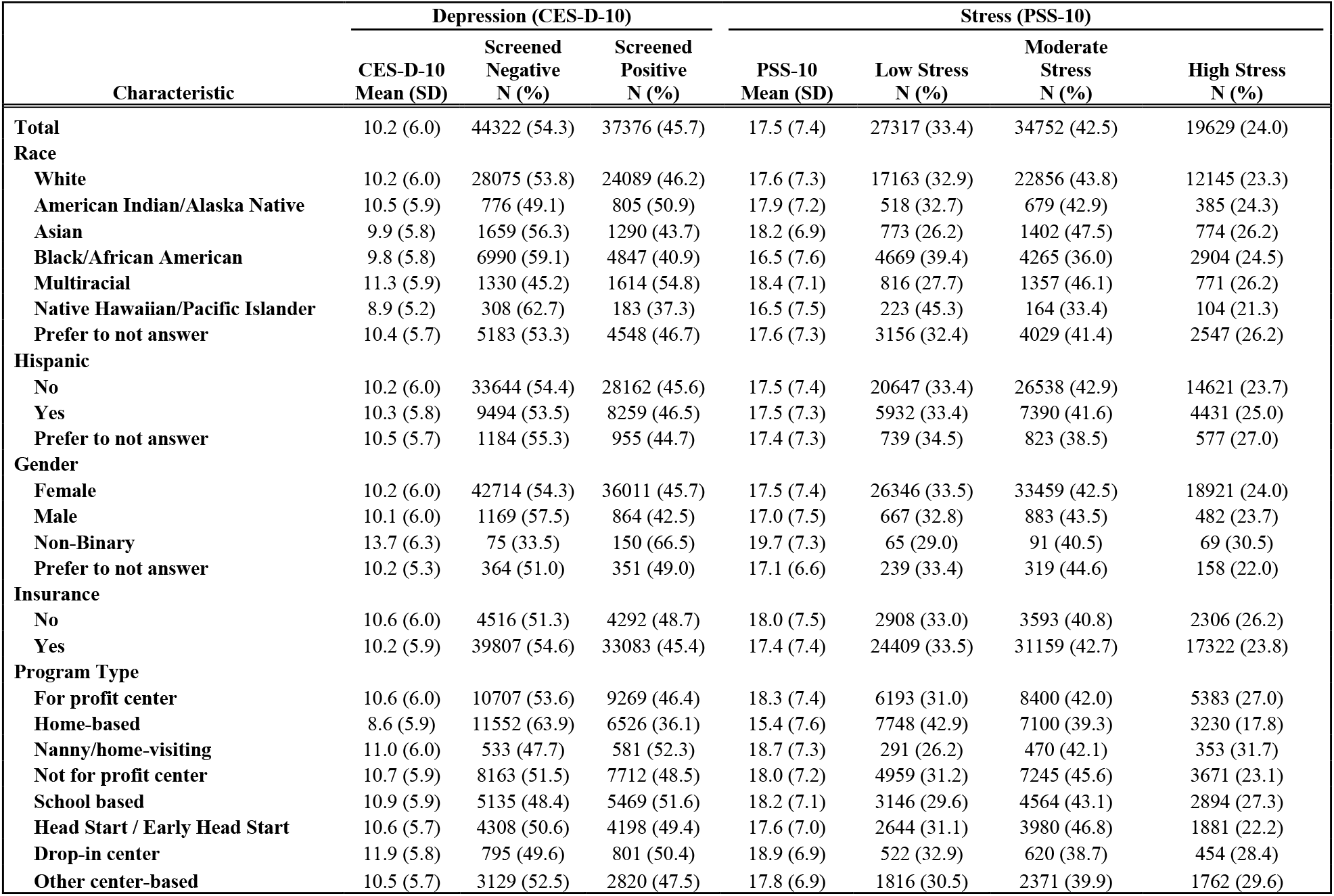
Prevalence of Mental Health Outcomes by Sociodemographic Characteristics.

| Characteristic | Depression (CES-D-10) |  |  | Stress (PSS-10) |  |  |  |
| --- | --- | --- | --- | --- | --- | --- | --- |
|  | CES-D-10<br>Mean (SD) | Screened<br>Negative<br>N (%) | Screened<br>Positive<br>N (%) | PSS-10<br>Mean (SD) | Low Stress<br>N (%) | Moderate<br>Stress<br>N (%) | High Stress<br>N (%) |
| **Total** | 10.2 (6.0) | 44322 (54.3) | 37376 (45.7) | 17.5 (7.4) | 27317 (33.4) | 34752 (42.5) | 19629 (24.0) |
| **Race** |  |  |  |  |  |  |  |
| **White** | 10.2 (6.0) | 28075 (53.8) | 24089 (46.2) | 17.6 (7.3) | 17163 (32.9) | 22856 (43.8) | 12145 (23.3) |
| **American Indian/Alaska Native** | 10.5 (5.9) | 776 (49.1) | 805 (50.9) | 17.9 (7.2) | 518 (32.7) | 679 (42.9) | 385 (24.3) |
| **Asian** | 9.9 (5.8) | 1659 (56.3) | 1290 (43.7) | 18.2 (6.9) | 773 (26.2) | 1402 (47.5) | 774 (26.2) |
| **Black/African American** | 9.8 (5.8) | 6990 (59.1) | 4847 (40.9) | 16.5 (7.6) | 4669 (39.4) | 4265 (36.0) | 2904 (24.5) |
| **Multiracial** | 11.3 (5.9) | 1330 (45.2) | 1614 (54.8) | 18.4 (7.1) | 816 (27.7) | 1357 (46.1) | 771 (26.2) |
| **Native Hawaiian/Pacific Islander** | 8.9 (5.2) | 308 (62.7) | 183 (37.3) | 16.5 (7.5) | 223 (45.3) | 164 (33.4) | 104 (21.3) |
| **Prefer to not answer** | 10.4 (5.7) | 5183 (53.3) | 4548 (46.7) | 17.6 (7.3) | 3156 (32.4) | 4029 (41.4) | 2547 (26.2) |
| **Hispanic** |  |  |  |  |  |  |  |
| **No** | 10.2 (6.0) | 33644 (54.4) | 28162 (45.6) | 17.5 (7.4) | 20647 (33.4) | 26538 (42.9) | 14621 (23.7) |
| **Yes** | 10.3 (5.8) | 9494 (53.5) | 8259 (46.5) | 17.5 (7.3) | 5932 (33.4) | 7390 (41.6) | 4431 (25.0) |
| **Prefer to not answer** | 10.5 (5.7) | 1184 (55.3) | 955 (44.7) | 17.4 (7.3) | 739 (34.5) | 823 (38.5) | 577 (27.0) |
| **Gender** |  |  |  |  |  |  |  |
| **Female** | 10.2 (6.0) | 42714 (54.3) | 36011 (45.7) | 17.5 (7.4) | 26346 (33.5) | 33459 (42.5) | 18921 (24.0) |
| **Male** | 10.1 (6.0) | 1169 (57.5) | 864 (42.5) | 17.0 (7.5) | 667 (32.8) | 883 (43.5) | 482 (23.7) |
| **Non-Binary** | 13.7 (6.3) | 75 (33.5) | 150 (66.5) | 19.7 (7.3) | 65 (29.0) | 91 (40.5) | 69 (30.5) |
| **Prefer to not answer** | 10.2 (5.3) | 364 (51.0) | 351 (49.0) | 17.1 (6.6) | 239 (33.4) | 319 (44.6) | 158 (22.0) |
| **Insurance** |  |  |  |  |  |  |  |
| **No** | 10.6 (6.0) | 4516 (51.3) | 4292 (48.7) | 18.0 (7.5) | 2908 (33.0) | 3593 (40.8) | 2306 (26.2) |
| **Yes** | 10.2 (5.9) | 39807 (54.6) | 33083 (45.4) | 17.4 (7.4) | 24409 (33.5) | 31159 (42.7) | 17322 (23.8) |
| **Program Type** |  |  |  |  |  |  |  |
| **For profit center** | 10.6 (6.0) | 10707 (53.6) | 9269 (46.4) | 18.3 (7.4) | 6193 (31.0) | 8400 (42.0) | 5383 (27.0) |
| **Home-based** | 8.6 (5.9) | 11552 (63.9) | 6526 (36.1) | 15.4 (7.6) | 7748 (42.9) | 7100 (39.3) | 3230 (17.8) |
| **Nanny/home-visiting** | 11.0 (6.0) | 533 (47.7) | 581 (52.3) | 18.7 (7.3) | 291 (26.2) | 470 (42.1) | 353 (31.7) |
| **Not for profit center** | 10.7 (5.9) | 8163 (51.5) | 7712 (48.5) | 18.0 (7.2) | 4959 (31.2) | 7245 (45.6) | 3671 (23.1) |
| **School based** | 10.9 (5.9) | 5135 (48.4) | 5469 (51.6) | 18.2 (7.1) | 3146 (29.6) | 4564 (43.1) | 2894 (27.3) |
| **Head Start / Early Head Start** | 10.6 (5.7) | 4308 (50.6) | 4198 (49.4) | 17.6 (7.0) | 2644 (31.1) | 3980 (46.8) | 1881 (22.2) |
| **Drop-in center** | 11.9 (5.8) | 795 (49.6) | 801 (50.4) | 18.9 (6.9) | 522 (32.9) | 620 (38.7) | 454 (28.4) |
| **Other center-based** | 10.5 (5.7) | 3129 (52.5) | 2820 (47.5) | 17.8 (6.9) | 1816 (30.5) | 2371 (39.9) | 1762 (29.6) |

**Table 4:** Linear Regression of CES-D-10, PSS-10, and Sociodemographic Characteristics.

| Characteristic | Depressive Symptoms (CES-D 10) |  | Stress (PSS-10) |  |
| --- | --- | --- | --- | --- |
| | $\beta$ (95% CI) | Std. Error | $\beta$ (95% CI) | Std. Error |
| **Age** | **-0.11 (-0.16 to -0.05)** | 0.03 | **-0.07 (-0.11 to -0.03)** | 0.02 |
| **Race** |  |  |  |  |
| **White** |  | *Reference* |  |  |
| **American Indian/Alaska Native** | 0.28 (-1.60 to 2.15) | 0.90 | 0.14 (-1.53 to 1.82) | 0.81 |
| **Asian** | 0.53 (-2.52 to 3.57) | 1.45 | -0.39 (-2.34 to 1.55) | 0.93 |
| **Black/African American** | -0.83 (-2.17 to 0.51) | 0.64 | -0.26 (-1.18 to 0.66) | 0.44 |
| **Multiracial** | 0.11 (-1.91 to 2.13) | 0.97 | 0.58 (-1.29 to 2.46) | 0.90 |
| **Native Hawaiian/Pacific Islander** | -0.60 (-5.21 to 4.01) | 2.21 | -1.09 (-4.59 to 2.41) | 1.68 |
| **Prefer to not answer** | 0.16 (-1.83 to 2.16) | 0.95 | 0.28 (-1.07 to 1.63) | 0.65 |
| **Hispanic** |  |  |  |  |
| **No** |  | *Reference* |  |  |
| **Yes** | -0.46 (-1.67 to 0.75) | 0.58 | -0.33 (-1.50 to 0.84) | 0.56 |
| **Prefer to not answer** | -0.16 (-2.46 to 2.13) | 1.10 | 0.08 (-2.62 to 2.79) | 1.29 |
| **Gender** |  |  |  |  |
| **Female** |  | *Reference* |  |  |
| **Male** | -0.95 (-3.52 to 1.61) | 1.23 | -0.45 (-2.29 to 1.38) | 0.88 |
| **Non-Binary** | 0.97 (-5.53 to 7.47) | 3.11 | 2.61 (0.26 to 4.95) | 1.14 |
| **Prefer to not answer** | -0.42 (-6.37 to 5.53) | 2.84 | -0.27 (-5.00 to 4.47) | 2.26 |
| **Insurance** |  |  |  |  |
| **Yes** |  | *Reference* |  |  |
| **No** | 0.26 (-1.49 to 2.01) | 0.83 | 0.27 (-1.09 to 1.63) | 0.65 |
| **Program Type** |  |  |  |  |
| **For profit center** |  | *Reference* |  |  |
| **Home-based** | **-2.30 (-3.89 to -0.72)** | 0.76 | **-1.54 (-2.45 to -0.63)** | 0.43 |
| **Nanny/home-visiting** | -0.49 (-3.70 to 2.73) | 1.54 | -0.20 (-3.03 to 2.63) | 1.35 |
| **Not for profit center** | -0.02 (-1.18 to 1.13) | 0.55 | 0.30 (-0.47 to 1.07) | 0.37 |
| **School based** | -0.40 (-2.10 to 1.30) | 0.81 | 0.11 (-0.87 to 1.09) | 0.47 |
| **Head Start / Early Head Start** | -0.73 (-2.77 to 1.30) | 0.97 | -0.07 (-1.24 to 1.10) | 0.56 |
| **Drop-in center** | -0.27 (-4.06 to 3.52) | 1.81 | 0.73 (-2.15 to 3.61) | 1.38 |
| **Other center-based** | -0.68 (-2.58 to 1.22) | 0.91 | -0.13 (-1.59 to 1.33) | 0.70 |

#### Stress

Of the total sample, 33.4% (n = 27,317) of child care professionals reported low stress levels, 42.5% (n = 34,752) reported moderate stress levels, and 24.0% (n = 19,629) reported high stress levels, with a mean score of 17.5 (std = 7.4). The prevalence of various stress levels and mean PSS-10 scores across sociodemographic characteristics can be found in **Table 3**. For every one-year increase in age, on average, the PSS-10 sum score decreased (β = -0.07; 95% CI = -0.11 to - 0.03). Compared to participants who worked in for-profit centers, participants in home-based programs, reported lower PSS-10 sum scores (β = -1.54; 95% CI = -2.45 to -0.63), while controlling for other covariates (**Table 4**).

## Discussion

In the largest national study of the physical and mental health of U.S. child care professionals, child care professionals during the COVID-19 pandemic reported a significantly higher depression rate compared to pre-pandemic estimates, and asthma and stress levels much greater than the U.S. adult estimates. Additionally, race, ethnicity, and gender disparities were evidenced for physical health conditions of child care professionals, but not for mental health during the pandemic. Our findings highlight a need for effective supports for the overall wellbeing of this vulnerable, yet essential, workforce.

The depression rate for child care professionals (45.7%) two to three months into the COVID-19 pandemic was greater than estimates for child care professionals prior to the pandemic (16.0% to 36.1%),^21-25^ and notably greater than estimates for U.S. adults during the pandemic (27.8% to 32.8%).^29-31^ In terms of stress levels, about two thirds (66.5%) of child care professionals reported moderate (42.5%) or high (24.0%) stress levels, almost twice the estimate for U.S. adults during the pandemic.^32^ This rate of stress among child care professionals is greater than pre-pandemic rates for child care professionals,^12,20,33^ and similar to rates during the early months of the pandemic reported by others.^12^

Across 10 medical conditions identified by the CDC near the beginning of the pandemic as risk factors for COVID-19 complications,^39^ 26.2% reported one, 9.7% reported two, and 4.0% reported three or more. Asthma rates for child care professionals in this study (14.3%) were about 1.2 times the national average for U.S. women.^47^ Asthma is associated with low-socioeconomic status and is more prevalent in communities of color, with those living in low-income neighborhoods at increased exposure to indoor allergens and airborne toxins often due to inadequately ventilated housing.^48-50^ Additionally, child care centers are often poorly ventilated and have high levels of indoor air pollutants,^51-54^ suggesting the need for greater attention to air quality in child care facilities to protect the health of child care professionals and reduce their vulnerability to COVID-19 complications.^55^ In contrast to asthma, child care professionals’ rates for diabetes (6.5%), heart disease (4.9%), and obesity (19.8%) were below national rates for U.S. adult women.^26,27,56^

Race, ethnicity, and gender disparities were evidenced for physical health conditions of child care professionals, but not for mental health during the pandemic. Compared to White child care professionals, those who identified as either American Indian/Native Alaskan or Black/African-American were at increased odds for asthma, heart disease, and diabetes, and multiracial child care professionals were at increased odds of asthma, heart disease, and obesity. Both findings are consistent with the racial and ethnic disparities among U.S. adult women.^27,47,56^ Illustrating these disparities, 7.4% of American Indian/Native Alaskan child care professionals reported three or more chronic health conditions that place them at greater risk of COVID-19 complications, compared to 4.0% for child care professionals overall. Of the physical health conditions examined, diabetes showed the greatest level of disparities, with all racial groups (except multiracial) and Hispanic showing increased odds when compared to White child care professionals, which is consistent with the racial and ethnic disparities among U.S. adult women.^57^ Also, child care professionals who reported non-binary gender identity were at increased odds of both asthma and heart disease.

In terms of child care setting, professionals working in the federally-funded Head Start or Early Head Start program were at greater odds for diabetes and obesity, compared to professionals working in for-profit child care centers, even when controlling for personal sociodemographic characteristics. Child care professionals with asthma, diabetes, or obesity were more likely to have access to health insurance regardless of age or other sociodemographic characteristics, perhaps explainable by previous findings showing that individuals with medical insurance being more likely to use basic clinical services and therefore more likely to receive a primary care diagnosis and treatment.^58^

### Strengths and Limitations

The major strength of our study is that it is a large national sample weighted to representativeness, allowing robust estimates of U.S. child care professionals’ physical and mental health status and enough statistical power to explore subgroup conditions. The greatest methodological limitation is the sole reliance on self-reported information without medical or psychiatric examination to verify the reporting. Also, findings were obtained during the early months of the COVID-19 pandemic (May-June 2020), and the mental health impacts of the COVID-19 pandemic may have changed over time.

### Conclusions

Given the impacts of the pandemic on this essential workforce, efforts should be directed toward developing effective and scalable interventions for improving their physical and mental health and addressing stressors that may undermine their wellbeing, such as long hours, low wages, and high job demands that are associated with child care professional stress, burnout and turnover.^6,8^ Our findings emphasize the need to further examine the health behaviors of child care professionals, via mixed-methods research, to understand what health initiatives might improve their overall wellbeing.

## Data Availability

All data produced in the present study are available upon reasonable request to the authors.

## Contributors’ Statement Page

Mr. Elharake designed the study, conducted the literature search, contributed to data interpretation, and drafted the initial manuscript. Ms. Shafiq designed the study, analyzed the data, contributed to data interpretation, and drafted the initial manuscript. Dr. Cobanoglu analyzed the data, contributed to data interpretation, and contributed to critical revision of the manuscript. Dr. Malik designed the study, analyzed the data, contributed to data interpretation, and contributed to revision of the manuscript. Prof. Humphries, Prof. Murray, Dr. Patel, Mr. Wilkinson, Prof. Yildirim, Ms. Diaz, Ms. Rojas, Ms. Kuperwajs Cohen, Mr. Lee, and Prof. Reyes contributed to data interpretation and contributed to critical revision of the manuscript. Ms. Klotz led data acquisition and development of the online survey tool, contributed to data interpretation, and contributed to critical revision of the manuscript. Prof. Omer designed the study, contributed to the analytic approach, contributed to data interpretation, and contributed to critical revision of the manuscript. Prof. Gilliam is the senior author who conceptualized the study, designed the study, conducted the literature search, was involved in aspects of data collection and analysis, and contributed to critical revision of the manuscript. All authors approved the final manuscript as submitted and agree to be accountable for all aspects of the work.

All authors are members of the Yale Children and Adults Research in Early Education Study (Yale CARES) team.

## Acknowledgements

This study was supported by The Andrew & Julie Klingenstein Family Fund, Esther A. & Joseph Klingenstein Fund, Heising-Simons Foundation, W.K. Kellogg Foundation, Foundation for Child Development, Early Educator Investment Collaborative, Scholastic Inc, Yale Institute for Global Health, and Tobin Center for Economic Policy at Yale University. Invaluable assistance with obtaining child care provider contact information was provided by the National Workforce Registry Alliance (and its network of state child care workforce registries), Child Care Aware of America, and National Association for the Education of Young Children. Drs. Amalia Londoño Tobón and Adrián Cerezo Caballero provided Spanish translations and back translations of the survey measures and recruitment information. Alicia Alonso, Catherine Chang, Renee Dauerman, Stella FitzGerald, Harleen Kaur, Emma Knight, and Helen Mooney provided assistance in qualitative data categorization of respondent comments.

## Supplementary Online Content

**eMethods**. Missing Data Analysis and Treatment

## eMethods

### Missing Data Analysis and Treatment

Missing data analysis included visual examination of missing data patterns and descriptive measures of missing values in the data. The chronic diseases and health conditions had 9.7% missing values, while CES-D-10 and PSS-10 scores had 36.1% and 37.3% missing, respectively. Missingness in covariates ranged between 0.5% and 16%. Little’s Missing Completely at Random (MCAR) test was used to examine the missing data mechanism, and results suggested that the data are not MCAR (χ^2^(9) = 1515.807, p < 0.001).^1^ Multiple Imputation (MI) was used to address missingness.^2^ Variables used for imputation included race, ethnicity, gender, age, child care program type, and access to medical insurance, and all the outcome variables i.e., CES-D-10, PSS-10, diabetes, heart disease, asthma, and obesity. Weight is also incorporated to fit imputation models. To ensure the precision and replicability of point estimates, we imputed 20 datasets using the Fully Conditional Specification Imputation Method,^3^ with pooled results from 20 datasets are reported in this paper.

---

**eTable 1:**
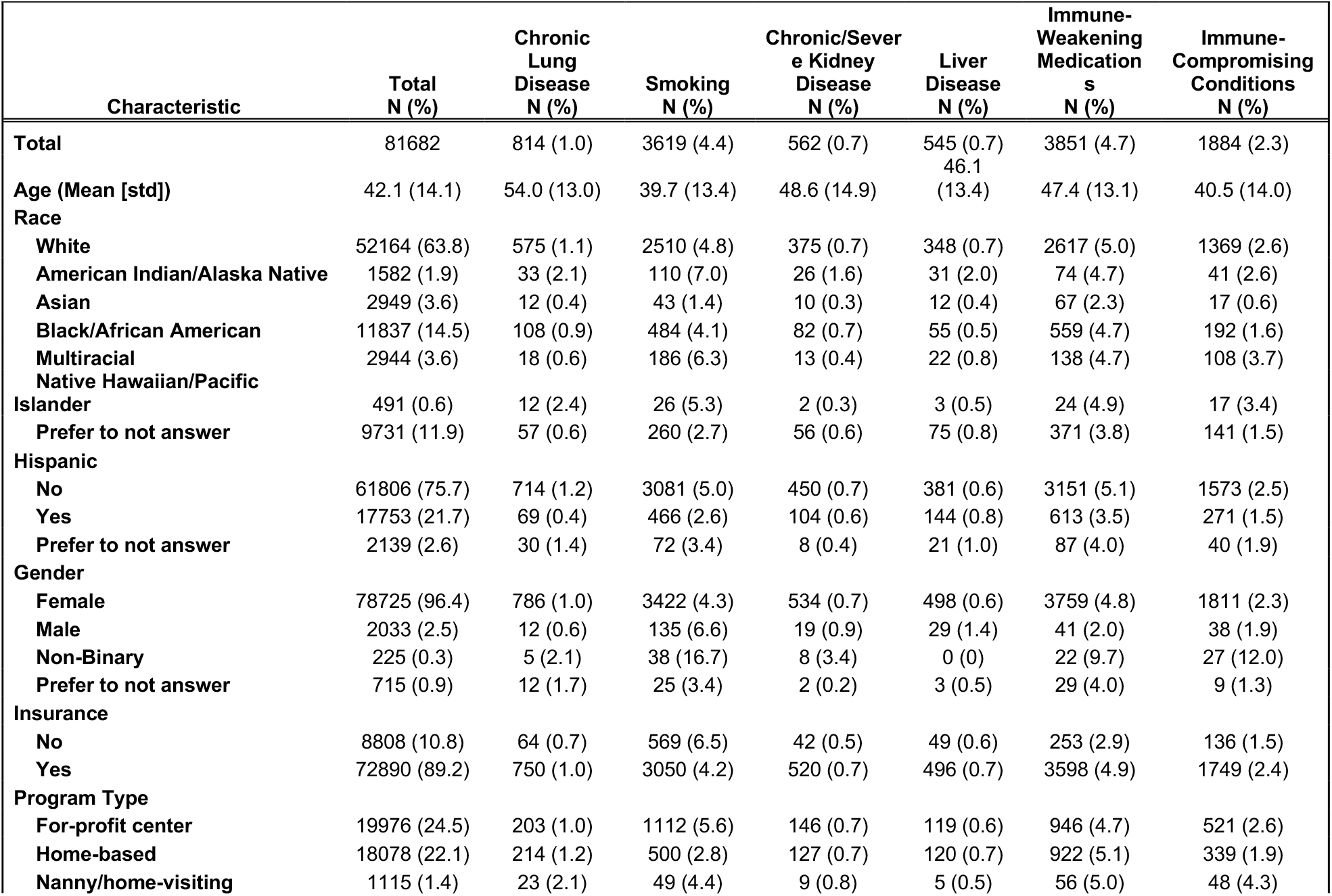
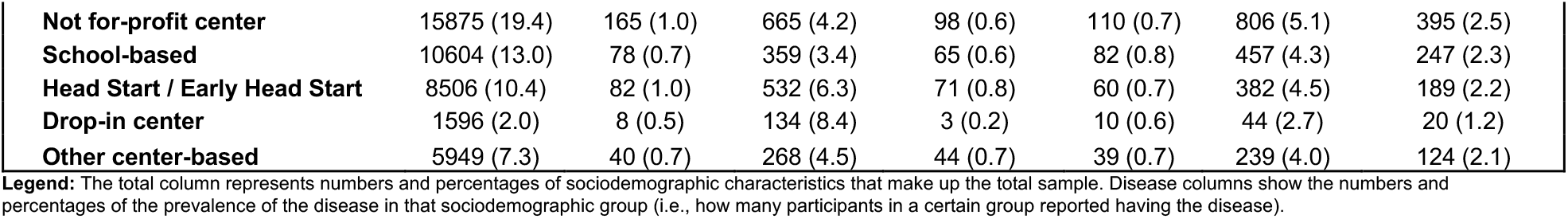
Prevalence of the Remaining Six Physical Health Outcomes by Sociodemographic Characteristics.

**eTable 2:** Number of Chronic Diseases and Health Conditions by Sociodemographic Characteristics.

| Characteristic | Number of Chronic Diseases of Health Conditions |  |  |  |
| --- | --- | --- | --- | --- |
|  | None<br>N (%) | One<br>N (%) | Two<br>N (%) | Three or<br>More<br>N (%) |
| **Total** | 49099 (60.1) | 21398 (26.2) | 7962 (9.7) | 3239 (4.0) |
| **Age (Mean [std])** | 41.1 (13.9) | 42.7 (14.1) | 44.5 (13.8) | 48.1 (13.7) |
| **Race** |  |  |  |  |
| **White** | 30874 (59.2) | 14009 (26.9) | 5174 (9.9) | 2107 (4.0) |
| **American Indian/Alaska Native** | 863 (54.6) | 421 (26.6) | 181 (11.4) | 117 (7.4) |
| **Asian** | 2221 (75.3) | 538 (18.3) | 138 (4.7) | 51 (1.7) |
| **Black/African American** | 6718 (56.8) | 3271 (27.6) | 1319 (11.1) | 530 (4.5) |
| **Multiracial** | 1561 (53.0) | 832 (28.3) | 406 (13.8) | 146 (4.9) |
| **Native Hawaiian/Pacific Islander** | 305 (62.1) | 113 (23.1) | 48 (9.7) | 25 (5.1) |
| **Prefer to not answer** | 6557 (67.4) | 2214 (22.8) | 697 (7.2) | 263 (2.7) |
| **Hispanic** |  |  |  |  |
| **No** | 35969 (58.2) | 16739 (27.1) | 6410 (10.4) | 2588 (4.3) |
| **Yes** | 11738 (66.1) | 4154 (23.4) | 1392 (7.8) | 468 (2.6) |
| **Prefer to not answer** | 1392 (65.1) | 505 (23.6) | 160 (7.5) | 82 (3.8) |
| **Gender** |  |  |  |  |
| **Female** | 47227 (60.0) | 20662 (26.2) | 7709 (9.8) | 3127 (4.0) |
| **Male** | 1300 (64.0) | 498 (24.5) | 169 (8.3) | 65 (3.2) |
| **Non-Binary** | 88 (39.2) | 80 (35.6) | 33 (14.9) | 23 (10.3) |
| **Prefer to not answer** | 484 (67.7) | 157 (21.9) | 51 (7.1) | 23 (3.3) |
| **Insurance** |  |  |  |  |
| **No** | 5654 (64.2) | 2212 (25.1) | 711 (8.1) | 231 (2.6) |
| **Yes** | 43445 (59.6) | 19186 (26.3) | 7251 (9.9) | 3008 (4.1) |
| **Program Type** |  |  |  |  |
| **For-profit center** | 11757 (58.9) | 5359 (26.8) | 2008 (10.1) | 852 (4.3) |
| **Home-based** | 11247 (62.2) | 4438 (24.5) | 1687 (9.3) | 706 (3.9) |
| **Nanny/home-visiting** | 717 (64.3) | 246 (22.1) | 103 (9.3) | 48 (4.3) |
| **Not for-profit center** | 9171 (57.8) | 4369 (27.5) | 1635 (10.3) | 699 (4.4) |
| **School-based** | 6755 (63.7) | 2692 (25.4) | 844 (8.0) | 313 (3.0) |
| **Head Start / Early Head Start** | 4783 (56.2) | 2369 (27.0) | 972 (11.4) | 381 (4.5) |
| **Drop-in center** | 1053 (66.0) | 374 (26.2) | 126 (7.9) | 43 (2.7) |
| **Other center-based** | 3617 (60.8) | 1549 (26.0) | 586 (9.8) | 197 (3.3) |

